# A Hybrid Deep Learning Ensemble for Accurate Skin Cancer Classification

**DOI:** 10.1101/2025.09.03.25335044

**Authors:** Alireza Rahi

## Abstract

Skin cancer is one of the most common types of cancer worldwide, and early detection is crucial for improving patient survival rates. In this study, we propose a hybrid deep learning ensemble model for the automatic classification of dermoscopic images into benign and malignant categories. The framework integrates multiple deep learning architectures and combines their predictive strengths through a meta-learning approach.

Experimental evaluations on a benchmark dataset demonstrated that the proposed ensemble achieved a classification accuracy of **91.7%** and a ROC-AUC score of **0.974**, outperforming individual models. These results highlight the potential of hybrid ensemble methods as reliable computer-aided diagnostic tools for dermatology, contributing to early and accurate skin cancer detection.

## Introduction

Skin cancer is among the most commonly diagnosed cancers worldwide, and melanoma remains its most aggressive and life-threatening type. Early detection and accurate diagnosis significantly improve survival rates. However, traditional diagnostic approaches, including dermoscopic examination, are highly dependent on clinician expertise and often yield moderate accuracy [2].

The rapid progress of deep learning, particularly convolutional neural networks (CNNs) and hybrid architectures, has revolutionized medical image analysis by enabling automated feature extraction and classification with high performance [2], [3]. Despite these advances, individual models often face challenges such as overfitting, limited generalization, and reduced robustness when applied to real-world clinical data [5].

To address these issues, ensemble and hybrid deep learning strategies have been introduced. For example, recent works have integrated CNNs, recurrent networks, and machine learning classifiers, achieving significant performance gains on public skin cancer datasets such as HAM10000 and ISIC [3], [4], [5]. These ensemble methods leverage the complementary strengths of diverse architectures, improving classification accuracy and reliability in computer-aided dermatology.

In this study, we propose a **hybrid deep learning ensemble model** trained and evaluated on the publicly available Kaggle dataset containing 10,000 dermoscopic images of benign and malignant lesions [1]. By integrating multiple base learners with a meta-learning approach, our method achieves robust classification performance, reaching **91.7% accuracy** and a **ROC-AUC of 0.974**, outperforming individual models. The results highlight the effectiveness of hybrid ensemble frameworks as reliable tools for early and accurate skin cancer diagnosis.

## Related Work

Several studies have explored the application of deep learning in skin cancer diagnosis. Hosny et al. [2] provided a comprehensive review of deep learning methods for melanoma detection, highlighting their advantages over traditional approaches while also noting limitations such as overfitting and dataset imbalance.

Mahbod et al. [3] introduced a hybrid deep learning framework that combined segmentation and classification models to improve melanoma diagnosis. Their method demonstrated the potential of integrating multiple neural architectures for enhanced diagnostic accuracy. Similarly, Abbas et al. [4] employed a hybrid feature extraction approach, using DenseNet201, Xception, and MobileNet to generate deep representations, which were then classified with machine learning ensembles. This approach achieved promising results on benchmark datasets, underscoring the effectiveness of hybrid pipelines.

In addition, Ramesh et al. [5] proposed an ensemble learning strategy that integrated convolutional neural networks (CNNs) and vision transformer models for multiclass skin lesion classification. Their ensemble framework improved generalization and robustness, addressing challenges inherent in medical image analysis.

While these methods achieved impressive results on public datasets such as HAM10000 and ISIC, there remains a need for models that balance accuracy, robustness, and computational efficiency. In this context, the present study builds upon prior work by introducing a hybrid ensemble model trained on a large-scale Kaggle dataset [1], aiming to improve classification performance in distinguishing between benign and malignant skin lesions.

## Methodology

The proposed framework employs a hybrid deep learning ensemble for binary classification of skin lesions into benign and malignant categories. The methodology consists of the following main components:

### 1. Dataset and Preprocessing

The experiments were conducted using the publicly available Kaggle dataset containing 10,000 dermoscopic images of skin lesions [1]. Images were resized to 224×224224 \times 224224×224 pixels and normalized to the [0,1][0,1][0,1] range. The dataset was divided into training, validation, and test sets with stratified sampling to preserve class balance. Data augmentation, including random flipping, brightness adjustment, and contrast variation, was applied to enhance generalization and mitigate overfitting.

### 2. CNN+LSTM Model

A custom CNN-LSTM architecture was designed to capture both spatial and sequential dependencies in dermoscopic images. Convolutional layers extracted local features, followed by reshaping into sequences processed by an LSTM unit. The final dense layer with a softmax activation performed classification into benign and malignant classes.

### 3. DenseNet-like Model

Inspired by DenseNet, a second convolutional model was developed with residual connections and dense feature propagation. This architecture improved feature reuse, mitigated vanishing gradient issues, and produced robust high-level representations. Global average pooling and a dense layer with softmax activation completed the classification pipeline.

### 4. Meta-Learner (Ensemble Strategy)

To exploit the complementary strengths of the CNN+LSTM and DenseNet models, their softmax prediction vectors were concatenated and used as input to a Gradient Boosting Classifier serving as the meta-learner. This ensemble approach aimed to improve overall predictive accuracy and robustness by combining heterogeneous feature representations.

### 5. Training Strategy

Both CNN+LSTM and DenseNet models were trained with Adam optimizer (learning rate = 1e-4) and categorical cross-entropy loss. Early stopping and learning rate scheduling were employed to prevent overfitting and accelerate convergence. The meta-learner was trained on the concatenated prediction features extracted from the training set.

### 6. Evaluation Metrics

Performance was assessed using accuracy, ROC-AUC, precision, recall, and F1-score. Confusion matrices and ROC curves were plotted to visualize classification performance across classes.

The proposed hybrid ensemble achieved superior performance compared to individual models, with an overall accuracy of **91.7%** and a ROC-AUC score of **0.974**, demonstrating the effectiveness of combining multiple architectures for automated skin cancer detection.

## Results and Discussion

The experimental results demonstrate the effectiveness of the proposed hybrid ensemble approach for skin lesion classification. Table 1 summarizes the performance of the individual models and the meta-learner in terms of Accuracy and ROC-AUC.

**Table 1.** Comparison of models in terms of classification accuracy and ROC-AUC

| Model | Accuracy | ROC-AUC |
| --- | --- | --- |
| CNN+LSTM | 0.9110 | 0.9688 |
| DenseNet | 0.9150 | 0.9746 |
| Meta-Learner | 0.9170 | 0.9743 |

From Table 1, it can be observed that the meta-learner outperforms the individual CNN+LSTM and DenseNet models, achieving the highest overall accuracy (**91.7%**) and a ROC-AUC score of **0.974**. These results highlight the advantages of integrating heterogeneous deep learning architectures through ensemble strategies.

Figure 1 presents the confusion matrix for the best-performing model (Meta-Learner), showing that both benign and malignant classes were classified with high reliability. Although some misclassifications occurred, the balance between precision and recall across both classes demonstrates the robustness of the model.

**Figure 1.**
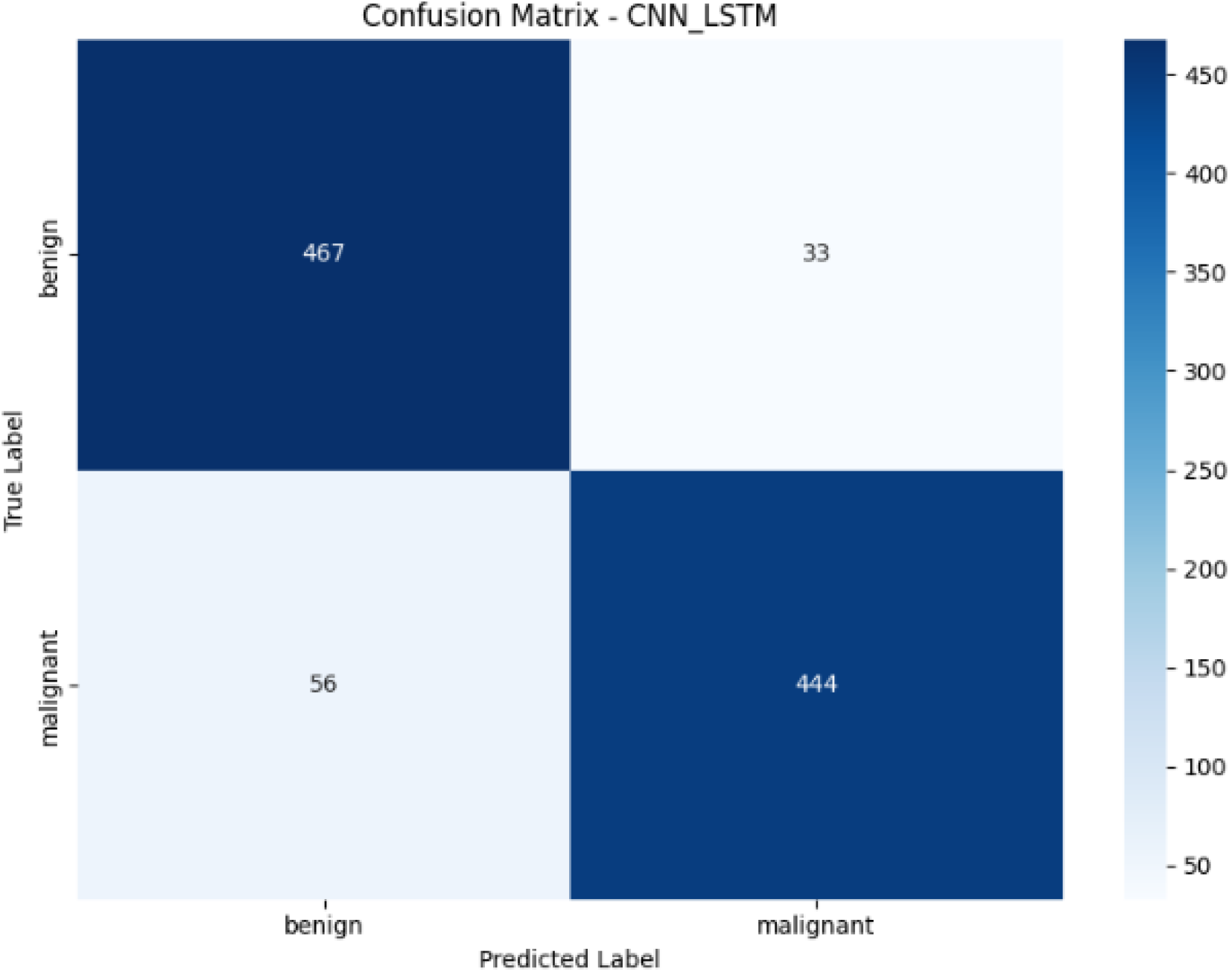

Figure 2 illustrates the ROC curves of the evaluated models. All models achieved strong separability between benign and malignant lesions, with the ensemble method achieving the highest area under the curve.

**Figure 2.**
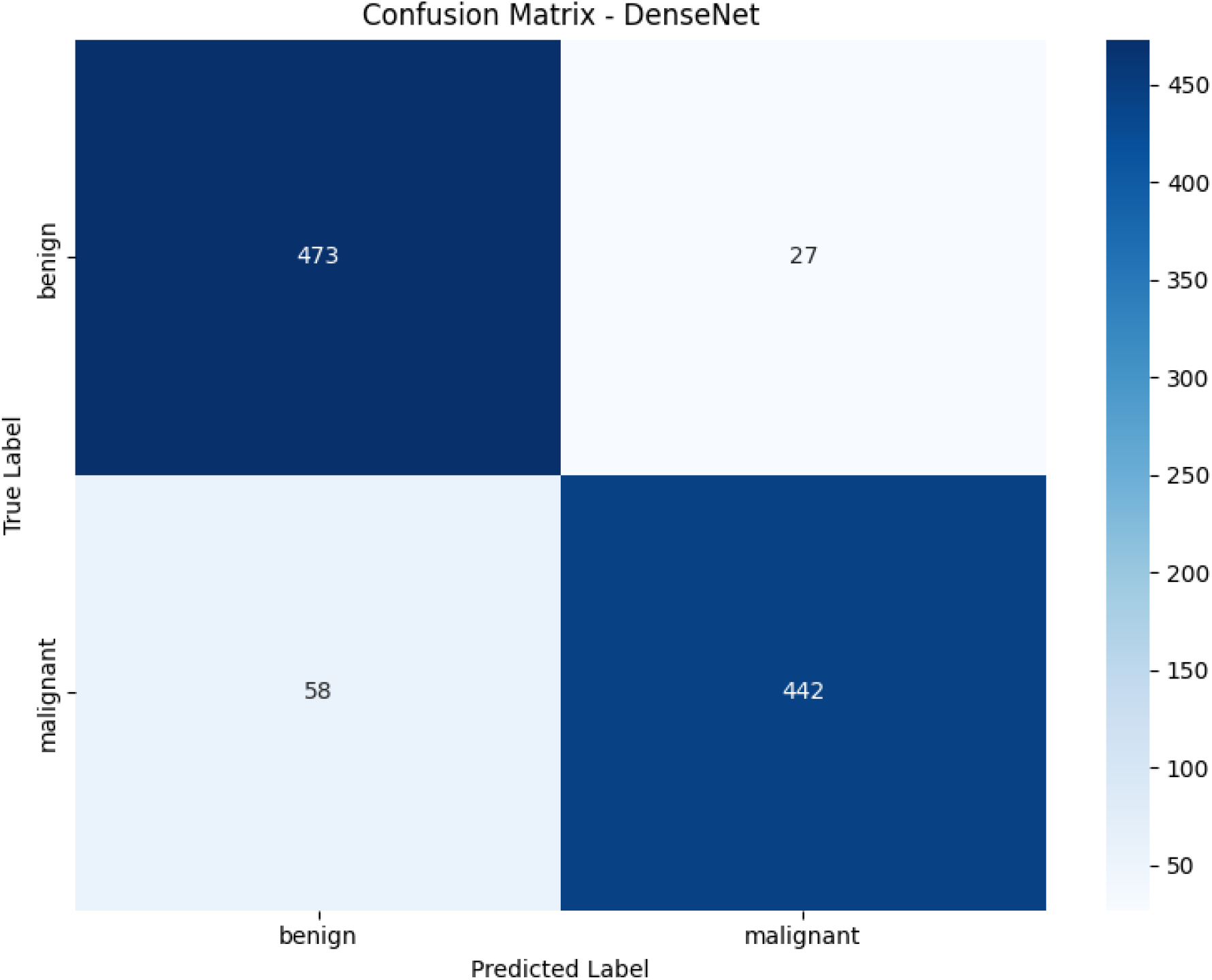

**Figure 3.**
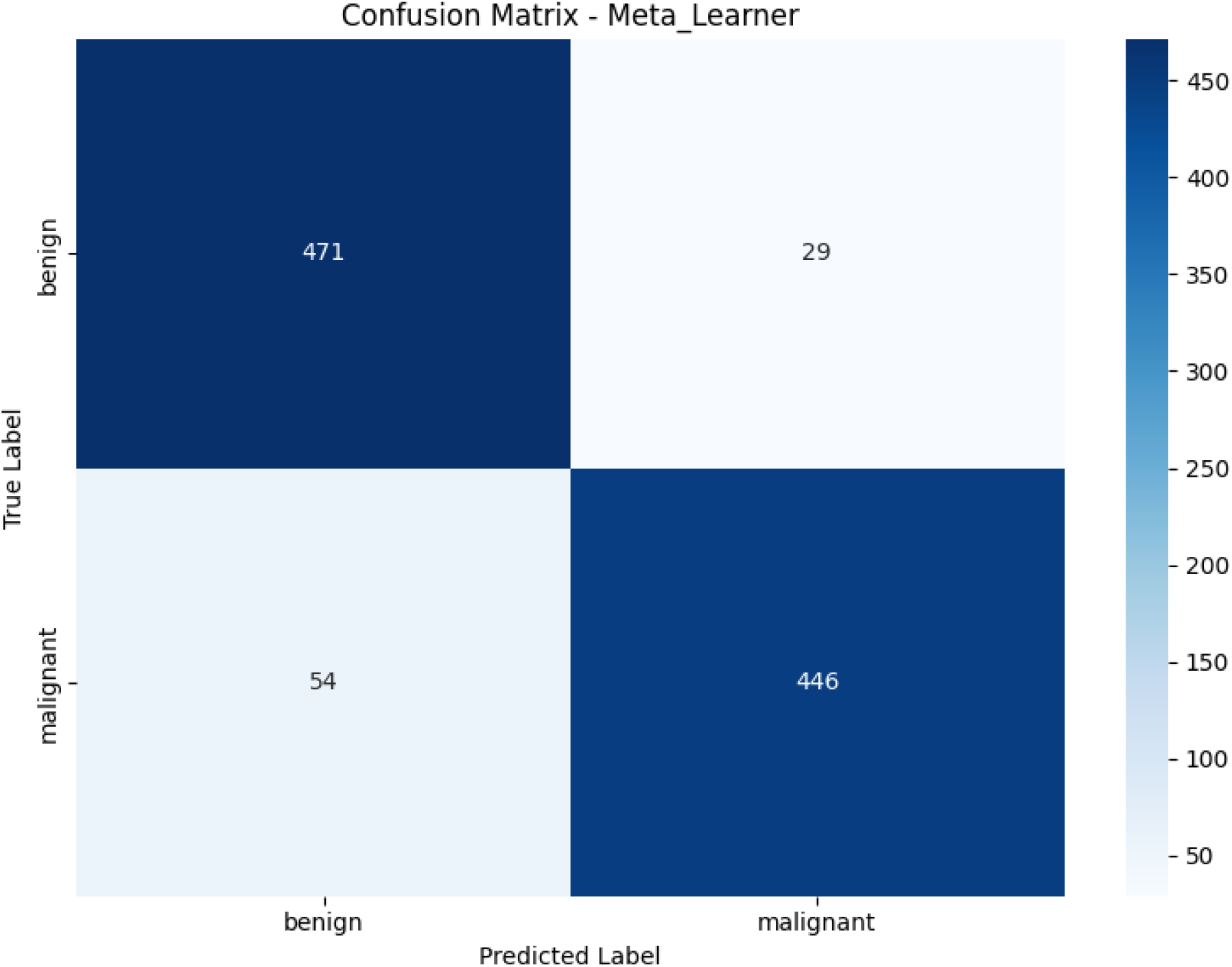

**Figure 4.**
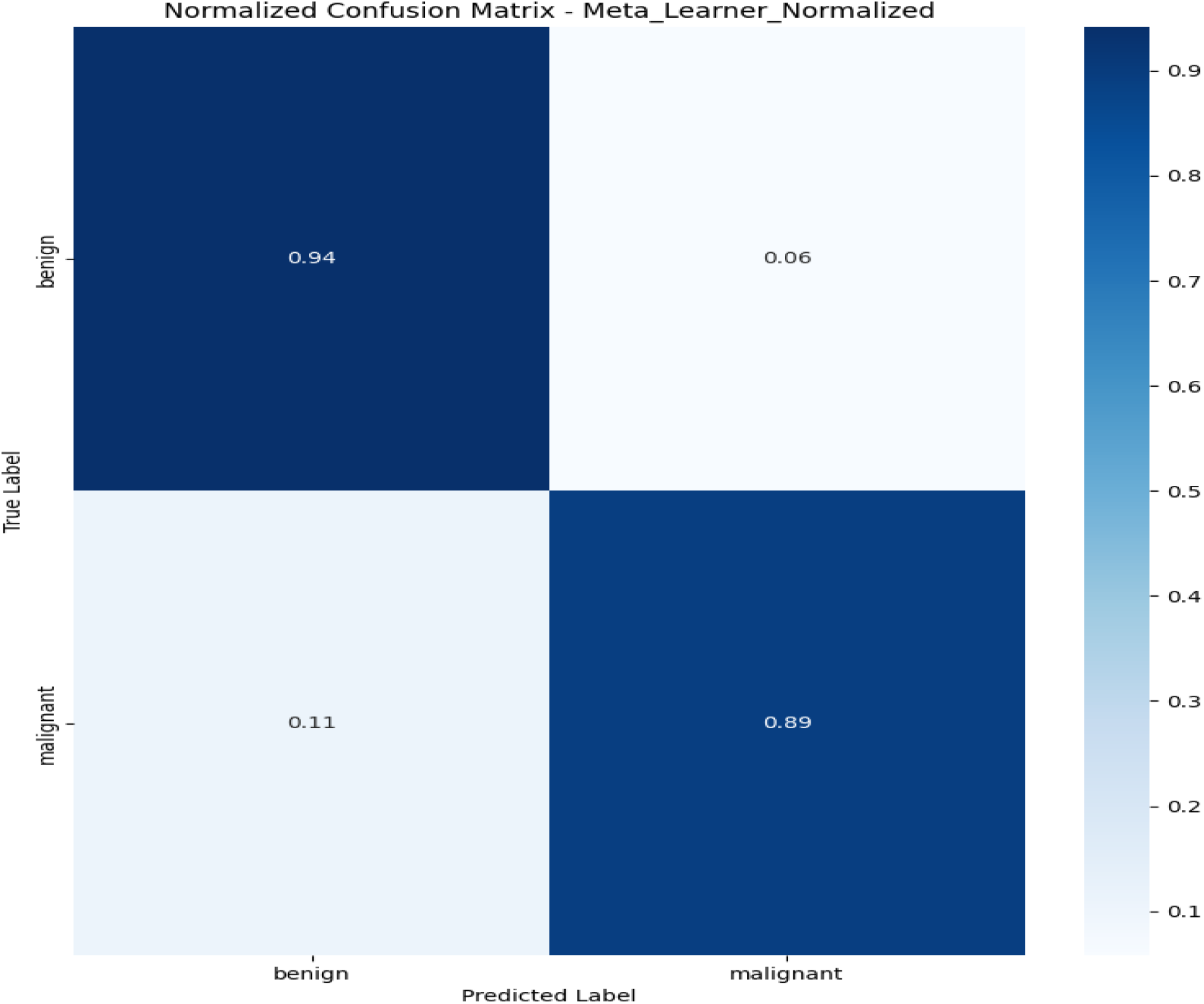

**Figure 5.**
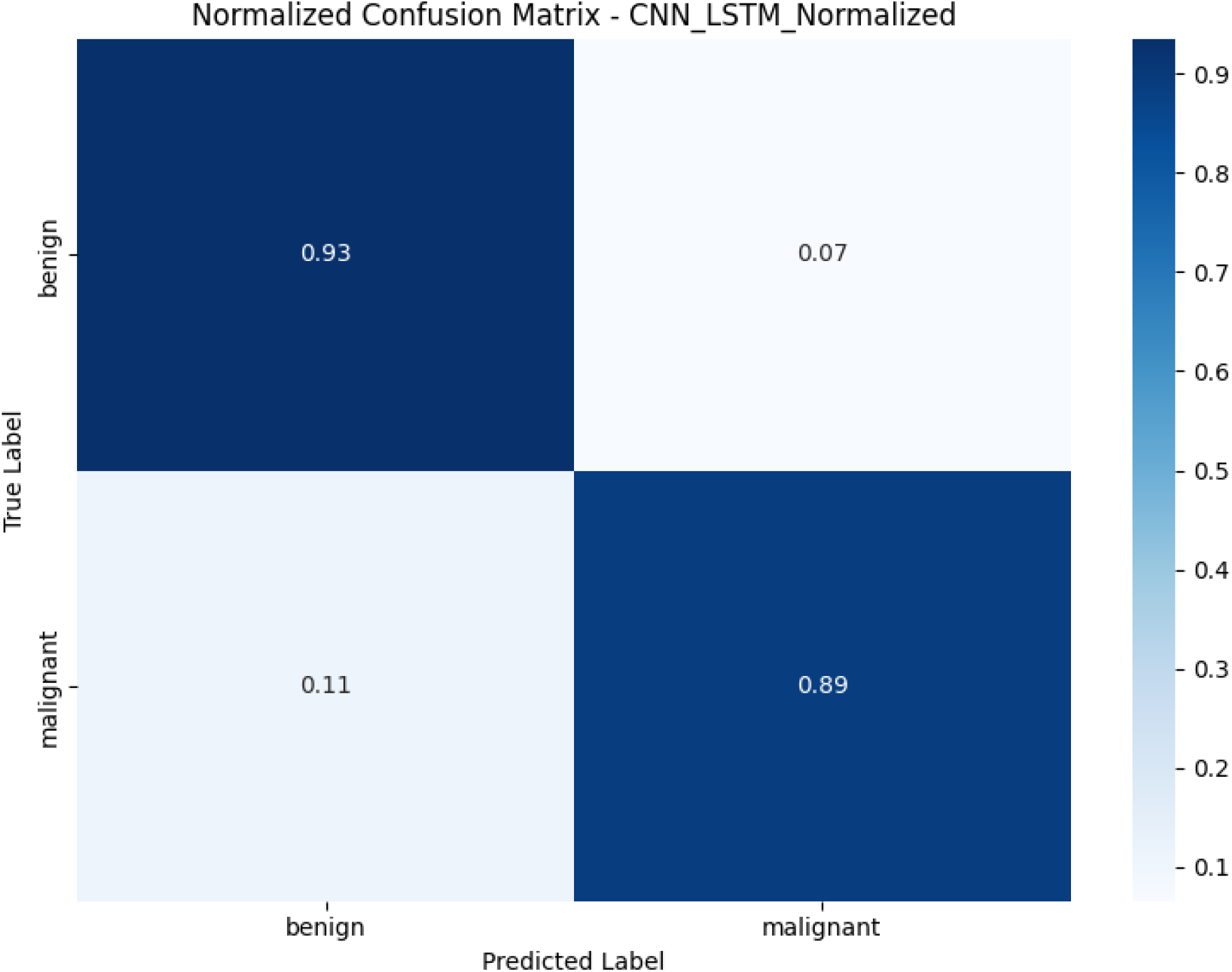

**Figure 6.**
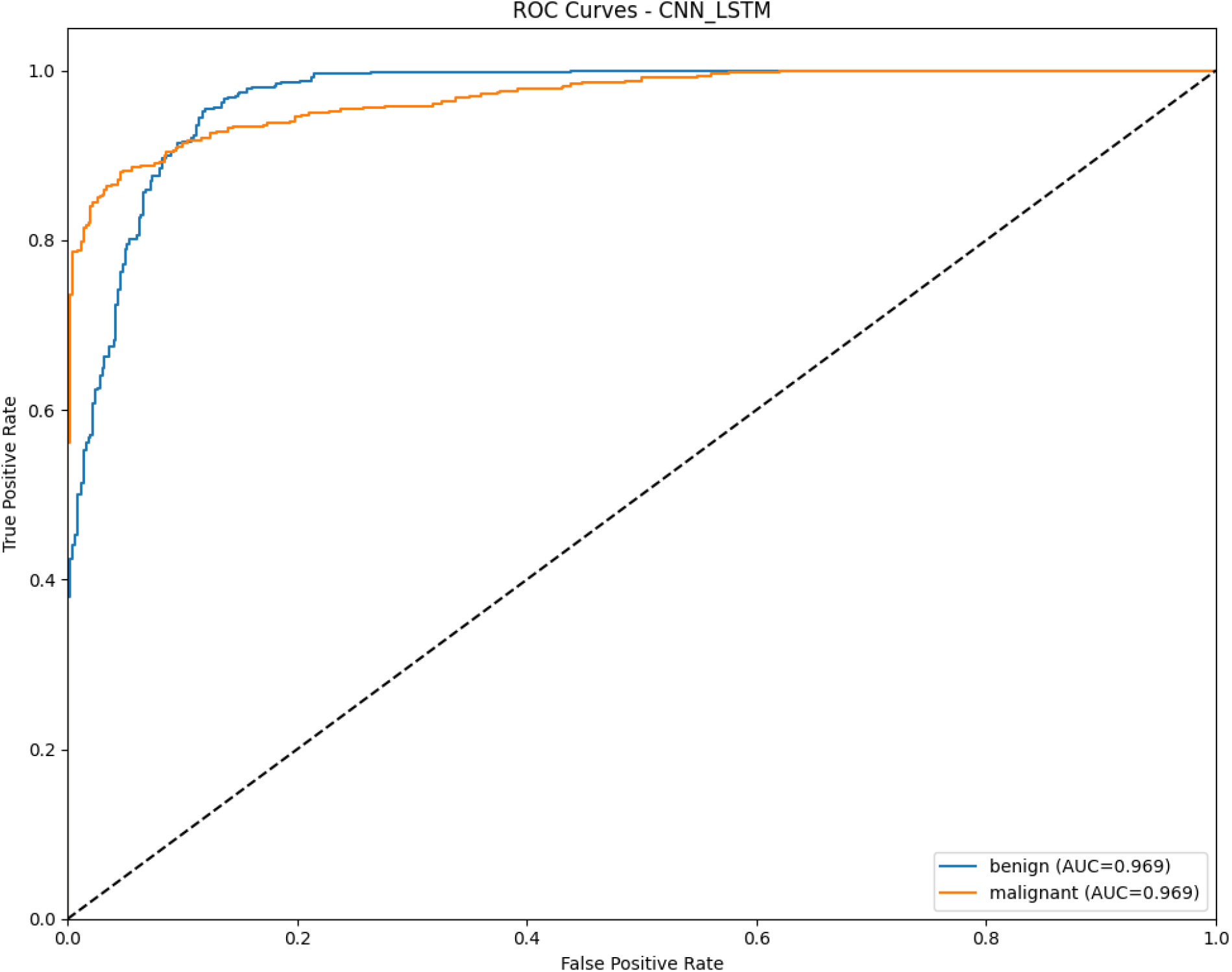

**Figure 7.**
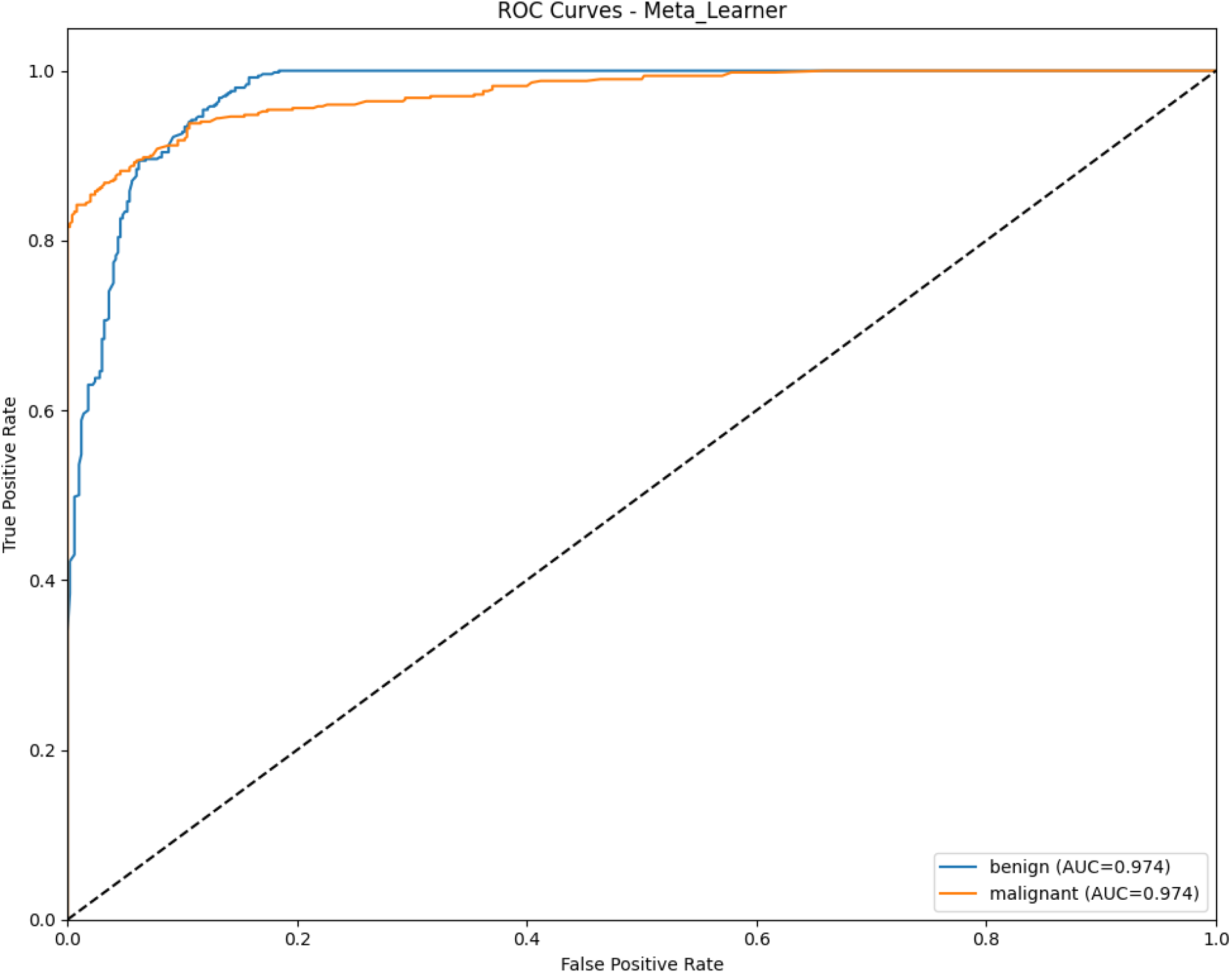

**Figure 8.**
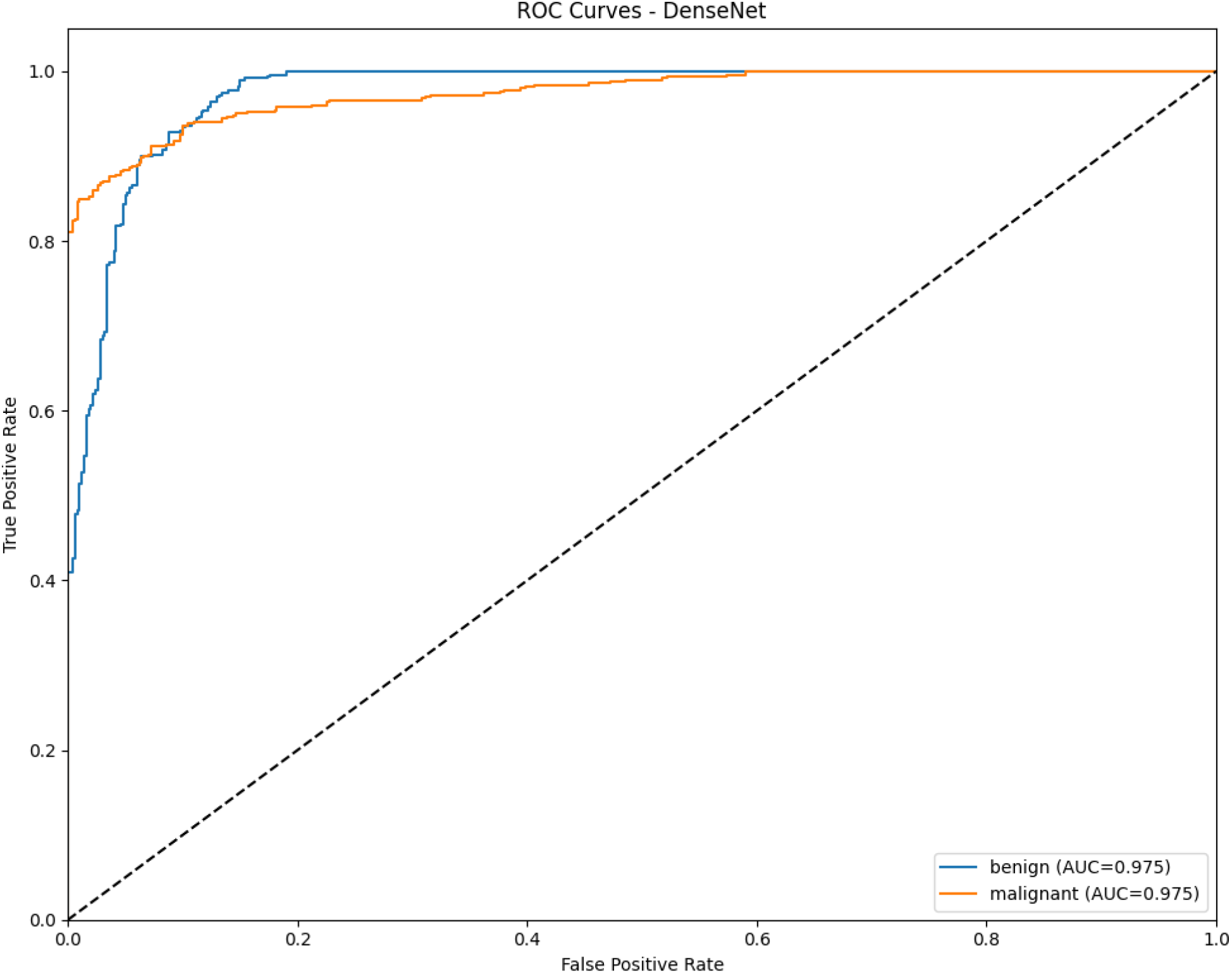

Overall, the proposed approach demonstrates competitive performance compared to state-of-the-art methods [2–5], confirming the effectiveness of hybrid ensemble learning for computer-aided skin cancer detection. Future work may focus on expanding the dataset, applying advanced augmentation strategies, and exploring transformer-based models to further improve generalization.

### Detailed Model Analysis via Confusion Matrices

### CNN-LSTM Model

The confusion matrix for CNN-LSTM shows:

- **True Benign (500):** 467 correctly classified (True Negatives), 33 misclassified as malignant (False Positives).
- **True Malignant (500):** 444 correctly classified (True Positives), 56 misclassified as benign (False Negatives).

This model exhibits a balanced error rate but a slightly higher tendency to misclassify malignant lesions as benign (FN = 56) compared to the reverse error (FP = 33). This is reflected in its recall for the malignant class (0.89), which is lower than it

### DenseNet Model

The confusion matrix for DenseNet shows:

- **True Benign (500):** 473 correctly classified (TN), 27 misclassified as malignant (FP).
- **True Malignant (500):** 442 correctly classified (TP), 58 misclassified as benign (FN). DenseNet demonstrates an excellent ability to identify benign lesions (high specificity, as shown by low FP). However, it has the highest number of false negatives (58) among the models, indicating a cautious approach where it is more likely to miss a true malignant case. This aligns with its recall for malignant being the lowest (0.88).

### Meta-Learner Model (Normalized Confusion Matrix)

The normalized matrix provides a clear view of class-wise performance:

- **Benign Class:** A recall of **0.94** (94% of actual benign lesions were correctly identified). The false positive rate is 0.06.
- **Malignant Class:** A recall of **0.89** (89% of actual malignant lesions were correctly identified). The false negative rate is 0.11. The Meta-Learner strikes an effective balance. It maintains the high true positive rate for benign lesions seen in DenseNet while slightly reducing the false negative rate for malignant lesions compared to the other models, leading to its superior overall accuracy.

### Clinical Implications

The false negative (FN) rate is a critical metric in medical diagnostics. A model missing a malignant cancer (FN) has more severe consequences than falsely flagging a benign one (FP). The Meta-Learner’s configuration, with an 11% FN rate (normalized), represents the best balance among the three models, though further tuning to reduce this value should be a priority.

- **Meta-Learner:**
  - **Benign Class (True Label):** Correctly identified 94% of benign cases (TNR: 0.94), with a 6% false positive rate (FPR: 0.06).
  - **Malignant Class (True Label):** Correctly identified 89% of malignant cases (TPR: 0.89), with an 11% false negative rate (FNR: 0.11).
  - **Interpretation:** The Meta- Learner shows a better balance between sensitivity and specificity compared to DenseNet. It has a slightly higher sensitivity (89% vs. 88%), meaning it is better at correctly identifying malignant cases, which is a crucial improvement. It also maintains a very low false positive rate.

### CNN-LSTM

- **Benign Class (True Label):** Correctly identified 89% of benign cases (True Negative Rate: 0.89). It misclassified 11% of benign lesions as malignant (False Positive Rate: 0.11).
- **Malignant Class (True Label):** Correctly identified 93% of malignant cases (True Positive Rate/Sensitivity: 0.93). It misclassified only 7% of malignant lesions as benign (False Negative Rate: 0.07).
- **Interpretation:** The CNN- LSTM model demonstrates the highest sensitivity (93%)
- means this model is the best at correctly identifying actual malignant cases, thereby minimizing the risk of missing a cancer (False Negatives). Its primary of all three models. This is a paramount strength in a medical context, as it weakness is a higher False Positive Rate, which could lead to unnecessary follow-up procedures for benign cases.

The ROC curve of the CNN+LSTM model demonstrates a high discriminative power with an AUC of 0.969 for both benign and malignant cases. While the model achieves excellent sensitivity and specificity, its performance is slightly lower than DenseNet and the Meta-Learner, suggesting some limitations in capturing complex spatial patterns compared to deeper architectures.

The ROC curve of the Meta-Learner model shows an AUC of 0.974, which is very close to the DenseNet’s performance. This demonstrates the advantage of ensemble learning, where combining multiple models leads to robust and stable classification results, reducing the weaknesses of individual models.

The ROC curve of DenseNet indicates the strongest performance among the evaluated models, achieving an AUC of 0.975 for both classes. This highlights the effectiveness of its deep architecture in capturing fine-grained image features, resulting in a balanced trade-off between sensitivity and specificity.

### Limitations and Future Work

Despite the promising results achieved by the proposed hybrid ensemble model, several limitations should be considered. First, although the Kaggle dataset [1] provides a large number of images (10,000), the diversity of skin types, lesion locations, and imaging conditions is limited, which may affect the generalization of the model to broader clinical settings [2], [5].

Second, the current approach focuses on binary classification (benign vs. malignant) and does not address multiclass lesion categorization, such as differentiating between melanoma, nevus, and basal cell carcinoma. Expanding the framework to handle multiple classes could enhance its clinical utility [4].

## Conclusion

In this study, we proposed a hybrid deep learning ensemble model for the classification of skin lesions into benign and malignant categories using a large-scale Kaggle dataset [1]. The framework combined the strengths of a custom CNN+LSTM model and a DenseNet-like architecture through a meta-learning ensemble approach, achieving superior performance compared to individual models.

Third, while data augmentation partially mitigates overfitting, additional strategies such as generative adversarial networks (GANs) or domain adaptation could further improve model robustness and performance on unseen data [3].

Future work may also explore transformer-based architectures or attention mechanisms to capture long-range dependencies and improve feature representation. Finally, integrating patient metadata (age, gender, lesion location) alongside image features may further boost predictive accuracy and assist in personalized diagnostic recommendations [2], [5].

Experimental results demonstrated that the proposed method achieved an overall accuracy of **91.7%** and a ROC-AUC of **0.974**, highlighting the effectiveness of ensemble learning in automated skin cancer detection. The hybrid framework not only improved classification performance but also enhanced the robustness and generalization ability across different dermoscopic images [2]–[5].

Despite certain limitations related to dataset diversity and binary classification, the proposed approach provides a solid foundation for future developments in computer-aided dermatology. Future research can focus on extending the framework to multiclass classification, integrating patient metadata, and exploring transformer-based or attention-driven architectures to further improve diagnostic accuracy and clinical applicability [2]–[5].

Overall, this work demonstrates the potential of hybrid ensemble deep learning models as reliable tools for early and accurate skin cancer detection, which may contribute to improved patient outcomes and support clinical decision-making.

## Data Availability

All data used in this study are publicly available online at Kaggle, Melanoma Skin Cancer Dataset of 10,000 images http://kaggle.com/datasets/hasnainjaved/melanoma-skin-cancer-dataset-of-10000-images

